# COVID-19 could cause long term peripheral nerve demyelination and axonal loss: A One Year Prospective Cohort Study

**DOI:** 10.1101/2022.07.19.22277248

**Authors:** Qian Yang, Meng Zhang, Yanhui Lai, Xuzhao Liu, Fengshuang Liu, Hongmin Zang, Jinzhong Song, Na Li, Shuhua Cui, Wei Shao, Jiang Ma, Zhibo Wang, Ling Cui, Feng Sun, Yubin Zhao

## Abstract

**Background:** There is a lack of studies on large-sample, medium-, or long-term follow-up data of peripheral neuropathy (PNP) in the COVID-19 survivors. This study evaluated the characteristics and related risk factors of PNP in the medium- and long-term rehabilitation, which provided real-world study data for the complete recovery of COVID-19 patients.

**Methods:** This study was a prospective cohort study of the COVID-19 survivors. We collected data on baseline characteristics, symptoms at onset and after discharge during the 6-month and 12-month follow-up. Peripheral nerves were measured by electromyography and inducible potentiometer. We used multivariable logistic regression to analyze the influencing factors of PNP. Additionally, we compared the difference between the two measurements among the population who completed both measurements.

**Results:** 313 patients were included in the study and all of them underwent nerve conduction study. 67 patients completed two measurements at 6-month and 12-month follow-up. Commonly reported symptoms contained memory loss (86%), hair loss (28%), anxiety (24%), and sleep difficulties (24%). 232 patients (74%) were found with PNP, including 51 (16%) with mononeuropathy and 181 (58%) with generalized PNP. Patients with measurement at 12-month follow-up had a higher prevalence of generalized PNP (p=0.006). For pathological types, 64 (20%) patients had only axonal loss, 67 (21%) had only demyelination, and 101 (32%) had a mixed type. There was no significant difference in the prevalence of accompanying symptoms after discharge between the two groups with or without PNP. After adjustment, age was positively associated with PNP (OR=1.22 per 10-year increase of age, 95% CI, 1.05-1.41). Compared with less than the median amount of IgG at discharge, higher amount of IgG was associated with decreased risk of F-wave abnormality (OR=0.32, 95%CI, 0.11-0.82), but no significant difference in other types of PNP.

**Conclusions and Relevance:** SARS-CoV-2 could cause PNP in hospital survivors with COVID-19, which persisted and was associated with age, education, and IgG antibody at discharge, but had no significant correlation with symptoms after discharge.

## Introduction

With the spread of the COVID-19 epidemic in the world, some studies^1-3^ reported the abnormal manifestations of the central and peripheral nervous systems of COVID-19 survivors. However, most of the previous studies were case reports for the acute phase^4,5^. Based on previous studies^6,7^, peripheral neuropathy (PNP) in hospital survivors with COVID-19 may have consisting effects on their daily functions and work ability. However, there is still no long-term follow-up and further measurement of these patients. High-quality and large-sample studies of peripheral nervous system in hospital survivors with COVID-19 are still quite limited.

The purpose of our study was to use a nerve conduction study (NCS) to analyze the overall characteristics, location, pathological type, and risk factors of PNP among COVID-19 survivors at 6-month and 12-month after diagnosis, offering a deeper insight into the medium- and long-term effects of SARS-CoV-2 on the peripheral nervous system. The results of this study may help clinicians formulate clinical rehabilitation directions for COVID-19 survivors and get them back to regular work and life.

## Methods

### Study design and participants

In this prospective cohort study, we selected patients who were diagnosed with COVID-19 from December 13, 2020 to April 14, 2021 and were discharged from Shijiazhuang People’s Hospital after treatment and routine rehabilitation. This study was reviewed and approved by the institution ethics committee of Shijiazhuang People’s Hospital, and all patients who agreed to participate in the study signed the informed consent form.

Inclusion criteria: (1) patients aged 10-70 years old. (2) patients diagnosed with COVID-19 and discharged from hospital by meeting the discharge criteria in the Eighth Edition of the Chinese Clinical Guidance for COVID-19 Pneumonia Diagnosis and Treatment issued by the National Health Commission of China^8^. (3) patients who could cooperate with researchers to carry out various examinations.

Exclusion criteria: (1) patients who were bedridden or with limited mobility; (2) patients with previous mental disorders, immune system diseases, neuromuscular diseases, malignant tumors, diabetes, fractures, cervical spondylosis and lumbar spondylosis; (3) patients who were unable to cooperate with researchers.

All the enrolled patients received rehabilitation treatment for 2 to 4 weeks after discharge, including cardiopulmonary function recovery, psychological counseling and traditional Chinese medicine treatment.

### Data Collection

The clinical data of COVID-19 patients at acute phase were obtained by searching the electronic medical records, including demographic characteristics, accompanying symptoms at onset, comorbidity, treatment, and novel coronavirus (nCoV) immunoglobulin G (IgG) at discharge. All participants were classified as mild, moderate, severe and critically severe according to the Eighth Edition of the Chinese Clinical Guidance for COVID-19 Pneumonia Diagnosis and Treatment^8^.

### Follow-up visit

Follow-up visits were completed in the COVID-19 rehabilitation clinic. All participants were interviewed face-to-face by trained medical staff and completed the consultation and a series of questionnaires. Borg Rating Scale of Perceived Exertion was used to evaluate the degree of fatigue of the patients. The Patient Health Questionnaire was used to screen for depression, and the Generalized Anxiety Disorder scale was used to filter the anxiety symptoms. We also recorded symptoms after discharge such as fever, sore throat, cough, and others.

### Nerve Conduction Study (NCS)

The full-featured electromyography/evoked potential equipment was used to measure the enrolled patients, and all patients underwent NCS. NCS included motor nerve conduction measurement and sensory nerve conduction measurement^9^. The measurements were completed by a physician with a senior professional title of neurophysiology, according to the conventional methods provided before^10^. The procedures and criteria were shown in the eMethods in the Supplement.

Electrophysiological evidence of axonal loss was reduced amplitude of sensory nerve action potential or compound muscle action potential with normal or slightly reduced conduction velocity (CV) and evidence of demyelination was markedly slowed conduction velocity or prolonged distal latency^11^. Judgment can be made according to the standards recognized by the respective laboratories or other laboratories^9,10^. PNP was divided into mononeuropathy, generalized PNP (including mononeuritis multiplex and polyneuropathy) according to European Standardized Telematic tool to Evaluate Electrodiagnostic Methods^11^. Pathological type was divided into demyelination, axon loss and mixed types. F-wave abnormality was judged by prolonged F-wave latency or less than 60% occurrence rate of F-wave.

### Outcome

The main outcomes of this study were PNP (including mononeuropathy and generalized PNP), and F-wave abnormality. The secondary outcomes were prevalence of PNP, CV, and amplitude in the results of two measurements among patients with both measurements.

### Statistical analysis

Categorical variables were described by count (percentage), and continuous variables by mean ± standard deviation (SD) or median (interquartile range, IQR). χ^2^ or Fisher’s exact test was used to compare the proportions in different subgroups; t test or Mann-Whitney U test was used for continuous variables.

For the main outcome, dichotomous indexes were used to describe the overall prevalence of PNP, mononeuropathy, generalized PNP, F-wave abnormality and prevalence of different pathological classifications (demyelination, axonal loss, and a mixed type), different anatomical locations (proximal or distal), different side of the body (left, right, and both), as well as the prevalence of each nerve injured. Continuous variables, such as CV and amplitude, were presented as mean ± SD. A human body diagram described the location and pathological classification of PNP. To compare CV and amplitude between 6-month and 12-month follow-up, we used paired t test or Wilcoxon signed-rank test when appropriate.

We used univariable and multivariable adjusted logistic regression analyses to explore risk factors associated with PNP, mononeuropathy, generalized PNP, and F-wave abnormality based on previous studies^12,13^. For the association of nCoV IgG at discharge with outcome, age, sex, smoking, education, comorbidity, antiviral, corticosteroids, and length of hospital stay were adjusted. The variables were all included in the model for the associations of sex, antiviral, corticosteroid, and length of hospital stay with the outcome. For the association of education with the outcome, the variables mentioned above except for comorbidity were included. For the association of smoking with the outcome, the variables mentioned above except for comorbidity, IgG at discharge, and length of hospital stay, were included. The variables mentioned above except for IgG at discharge and length of hospital stay, were included for the association of comorbidity with the outcome. Only sex, cigarette smoking, and education were adjusted for the association of age with the outcome.

Statistical analyses were performed using R (version 4.1.3) and SPSS (version 26.0). Two-tailed p<0.05 were considered statistically significant.

## Results

332 participants were enrolled (161 in 6-month follow-up and 171 in 12-month follow-up) and received at least one NCS. We excluded 19 patients with blood sugar fluctuations during follow-up or without complete records. There were 313 participants included in the final analysis and 67 of them received both measurements in 6-month and 12-month visits (Figure 1).

**Figure 1:**
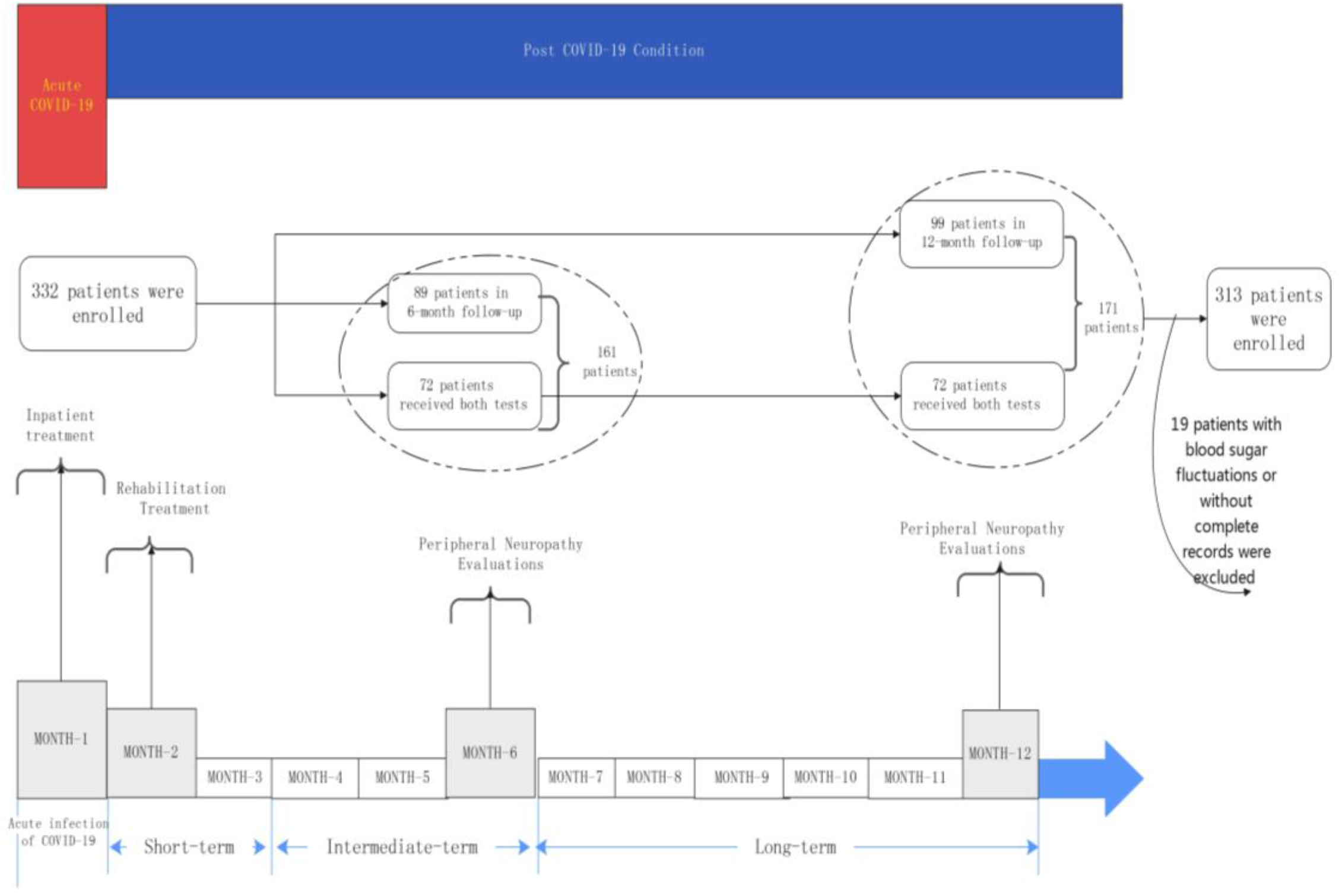
A framework and timeline of this study.

As table 1 showed, after admission, Median follow-up time was 350.0 days (IQR 165.0-357.0). The median age was 49.0 (IQR 33.0-58.0) years old and 191 (61%) were women. 96 (31%) patients were classified as mild, 200 (64%) as moderate, and 17 (5%) as severe or critically ill. The median hospital stay was 17.0 (IQR 14.0-23.0) days. 32 (10%) patients received antiviral therapy and 7 (2%) patients received corticosteroids therapy during hospital stay (Table 1).

**Table 1:** Characteristics of baseline of enrolled patients.

|  | **Total**<br>**(n=313)** | **No peripheral**<br>**neuropathy (n=81)** | **Peripheral**<br>**neuropathy (n=232)** | **p**<br>**value** |
| --- | --- | --- | --- | --- |
| Age, mean ± SD, years | 44.7 ± 17.5 | 39.8 ± 17.7 | 46.5 ± 17.1 | 0.003 |
| Age, median (IQR), years | 49.0 (33.0-58.0) | 43.0 (28.0-53.0) | 51.0 (34.8-59.0) | 0.002 |
| Sex |  |  |  | 0.12 |
| Male | 122 (39%) | 38 (47%) | 84 (36%) |  |
| Female | 191 (61%) | 43 (53%) | 148 (64%) |  |
| Smoking |  |  |  | 0.91 |
| Never-smoker | 283 (90%) | 74 (91%) | 209 (90%) |  |
| Current or former smoker | 30 (10%) | 7 (9%) | 23 (10%) |  |
| Education |  |  |  | 0.75 |
| Middle school or lower | 294 (94%) | 75 (93%) | 219 (94%) |  |
| College or higher | 19 (6%) | 6 (7%) | 13 (6%) |  |
| Disease severity |  |  |  | 0.25 |
| Mild | 96 (31%) | 29 (36%) | 67 (29%) |  |
| Moderate | 200 (64%) | 50 (62%) | 150 (65%) |  |
| Severe or critically ill | 17 (5%) | 2 (3%) | 15 (7%) |  |
| Symptoms and signs at disease onset |  |  |  |  |
| Fever | 64 (20%) | 16 (20%) | 48 (21%) | 0.98 |
| Cough | 70 (22%) | 18 (22%) | 52 (22%) | 0.99 |
| Dyspnea | 16 (5%) | 4 (5%) | 12 (5%) | 0.99 |
| Fatigue | 17 (5%) | 2 (3%) | 15 (7%) | 0.28 |
| Nausea or vomiting | 9 (3%) | 1 (1%) | 8 (3%) | 0.52 |
| Diarrhea | 5 (2%) | 0 (0%) | 5 (2%) | 0.41 |
| Myalgia | 8 (3%) | 1 (1%) | 7 (3%) | 0.64 |
| Chest tightness | 12 (4%) | 2 (3%) | 10 (4%) | 0.68 |
| Sputum production | 33 (11%) | 6 (7%) | 27 (12%) | 0.39 |
| Comorbidity | 78 (25%) | 17 (21%) | 61 (26%) | 0.42 |
| Hypertension | 67 (21%) | 15 (19%) | 52 (22%) | 0.56 |
| Heart diseases | 18 (6%) | 5 (6%) | 13 (6%) | 0.99 |
| Cerebrovascular diseases | 4 (1%) | 1 (1%) | 3 (1%) | 0.99 |
| Chronic obstructive pulmonary diseases | 5 (2%) | 1 (1%) | 4 (2%) | 0.99 |
| Antiviral therapy | 32 (10%) | 5 (6%) | 27 (12%) | 0.24 |
| Corticosteroids therapy | 7 (2%) | 2 (3%) | 5 (2%) | 0.99 |
| nCoV IgG at discharge, median (IQR), S/CO | 31.8 (12.2-120.2) | 23.1 (9.1-77.9) | 35.6 (13.0-130.8) | 0.08 |
| Time from admission to follow-up, median (IQR), days | 350.0 (165.0-357.0) | 349.0 (154.0-356.0) | 350.0 (324.5-357.0) | 0.14 |
| Length of hospital stay, median (IQR), days | 17.0 (14.0-23.0) | 17.0 (14.0-22.0) | 17.0 (14.0-23.0) | 0.48 |

Abbreviation: SD, standard deviation; IQR, interquartile range; nCoV IgG, novel coronavirus immunoglobulin G; S/CO=sample/cut off.

Among 311 patients, 232 (74%) patients had PNP. Patients with PNP were significantly older compared with those without it (51.0 (IQR 34.8-59.0) vs 43.0 (IQR 28.0-53.0)). We found no significant difference of sex, smoking, disease severity, and comorbidity between two groups (Table 1). Demographics and baseline characteristics of 67 patients with two measurements was shown in eTable 1 in the Supplement.

Figure 2 and eTable 2 in the Supplement shows the characteristics of PNP of 313 patients during follow-up. The results at 12-month follow-up were considered for patients who received two measurements. 232 (74%) patients had PNP, 55 (66%) for 6-month follow-up, and 177 (77%) for 12-month follow-up (p=0.08). 51 (16%) patients had mononeuropathy, 18 (22%) for 6-month follow-up and 33 (14%) for 12-month follow-up. 181 (58%) patients had generalized PNP, 37 (45%) for 6-month follow-up and 144 (63%) for 12-month follow-up (p=0.007). According to the prevalence from high to low, the nerve injuries at 6-month follow-up were as follows: 31 (37%) patients with median nerve injured, 24 (29%) with peroneal nerve injured, 19 (23%) with ulnar nerve injured, 12 (15%) with tibial nerve injured, and 9 (11%) with sural nerve injured. At 12-month follow-up, 104 (45%) patients were found with median nerve injured, 87 (38%) with peroneal nerve injured, 65 (28%) with tibial nerve injured, 47 (20%) with sural nerve injured, and 45 (20%) with ulnar nerve injured. Prevalence of tibial nerve injured at 12-month follow-up was significantly higher than that of 6-month follow-up (p=0.02).

**Figure 2:**
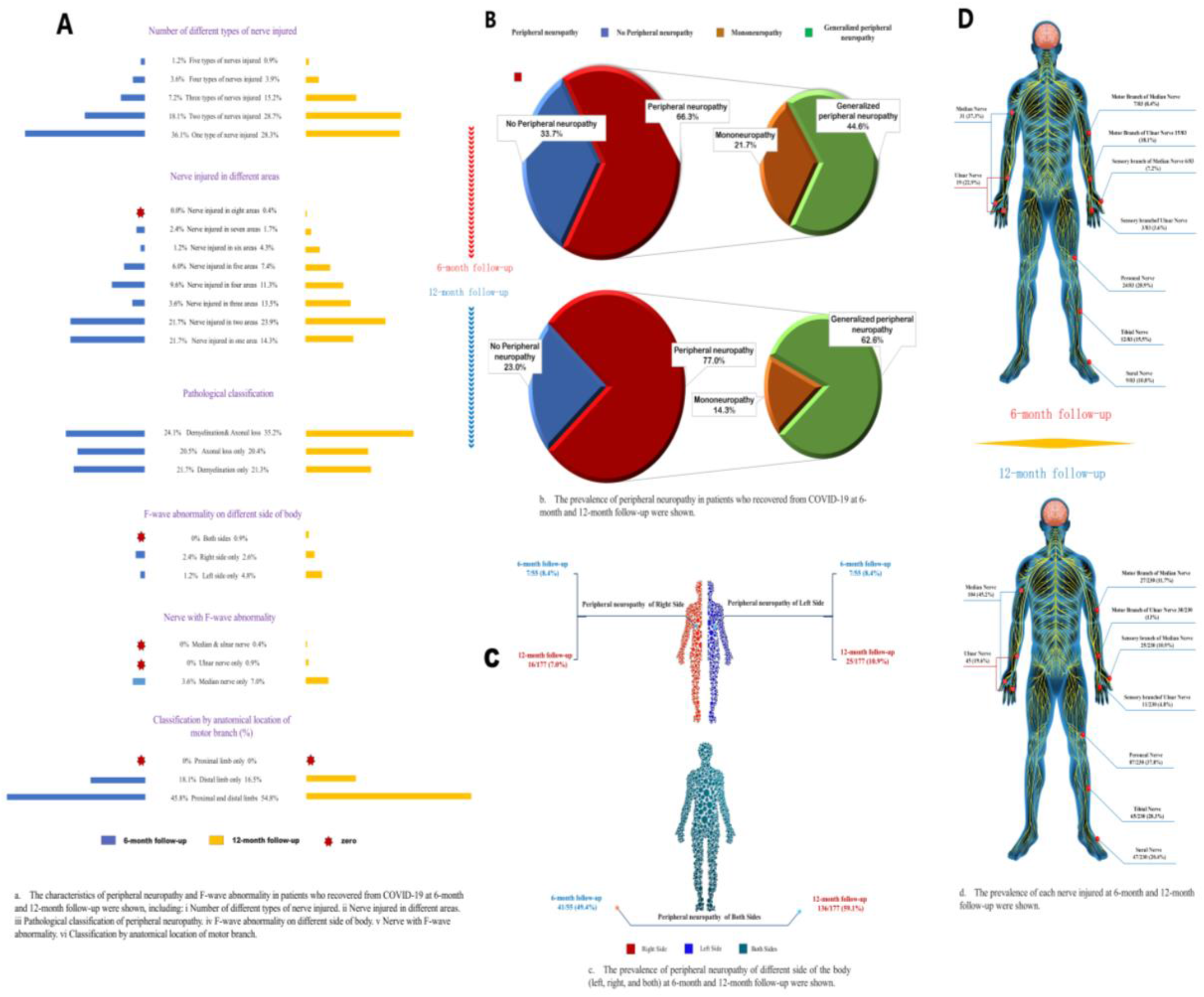
Prevalence and characteristics of different subtypes of peripheral neuropathy.

81 (26%) patients were found two types of nerves injured, among which the median nerve and the peroneal nerve were mainly injured at the same time. 41 (13%) patients were found with three types of nerves injured, 12 (4%) with four types, and 3 (1%) with five types (eTable 3 in the Supplement). In terms of pathological types, 64 (20%) patients only had axonal loss, 67 (21%) only had demyelinating lesions, and 101 (32%) had a mixed type. There was a significant difference in the pathological type distribution of peroneal nerve at 6-month and 12-month follow-up (p=0.019) and prevalence of demyelinating lesions decreased (6-month: 6%; 12-month: 1%), while rate of axonal loss increased (6-month: 21%; 12-month: 34%, eTable 4 in the Supplement). 32 (10%) had PNP only on the left side, 23 (7%) on the right side, and 177 (57%) with bilateral injury. Prevalence of only distal injury was 17% and 164 (52%) patients had both proximal and distal injuries. 22 (7%) patients had F-wave abnormality, and 19 (6%) had F-wave abnormality only in the median nerve (eTable 2).

The symptoms among patients with and without PNP at follow-up were shown in eTable 5 in the Supplement. Memory loss was the most commonly reported symptom. 269 (86%) patients presented memory loss, including 74 patients with PNP and 195 without PNP. Other common symptoms included hair loss (28%), anxiety (24%), sleep difficulties (24%), muscle weakness (21%), dry throat (21%), and fatigue (20%). There was no significant difference in any symptoms and signs between patients with and without PNP.

Univariable and multivariable logistic analyses showed similar results (eTable 6 and Figure 3). After adjustment, age was positively associated with PNP (odds ratio (OR)=1.22 per 10-year increase of age, 95% confidence interval (CI), 1.05-1.41) and generalized PNP (OR=1.16, 95% CI, 1.01-1.32). There was no significant association between age and F-wave abnormality (OR=1.00, 95% CI, 0.78-1.29). Compared with middle school or lower education, patients with college or higher education had a higher risk of mononeuropathy (OR=5.07, 95% CI, 1.80-13.90) and lower risk of generalized PNP (OR =0.27, 95% CI, 0.09-0.75). Compared with less than the median amount of nCoV IgG at discharge, higher amount of IgG was associated with decreased risk of F-wave abnormality (OR=0.32, 95%CI, 0.11-0.82), but no significant difference in other types of PNP (Figure 3).

**Figure 3:**
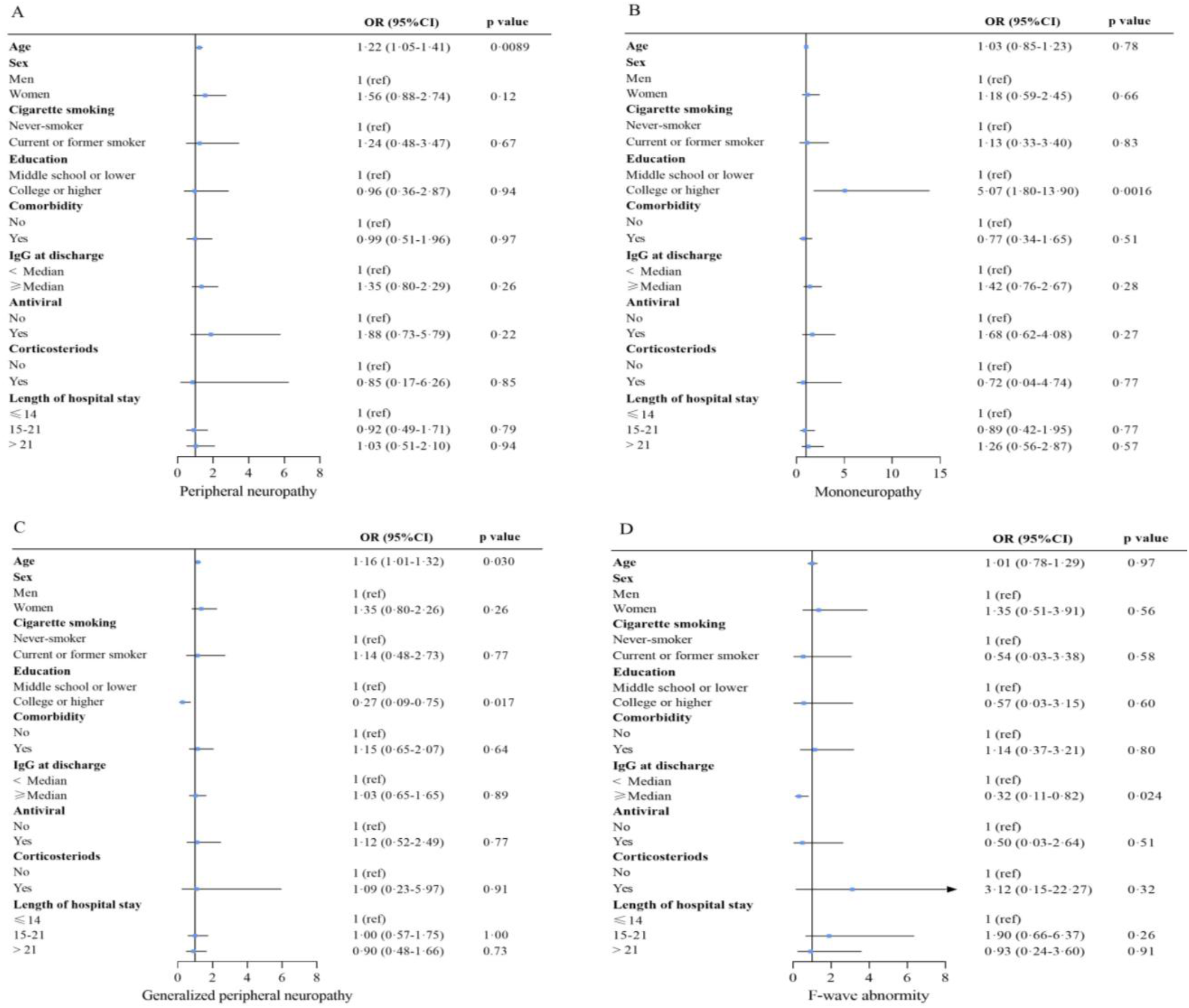
Risk factors associated with peripheral neuropathy (A), mononeuropathy (B), generalized peripheral neuropathy (C), and F-wave abnormality (D) at 6 or 12-month follow-up. For the association of nCoV IgG at discharge with outcome, age, sex, smoking, education, comorbidity, antiviral, corticosteroids, and length of hospital stay were adjusted. For the association of sex, antiviral, corticosteroid, and length of hospital stay with outcome, the aforementioned variables were all included in the model. For the association of education with outcome, the aforementioned variables except for comorbidity were included. For the association of smoking with outcome, the aforementioned variables except for comorbidity, IgG at discharge, and length of hospital stay were included. For the association of comorbidity with outcome, the aforementioned variables except for IgG at discharge and length of hospital stay were included. For the association of age with outcome, only sex, cigarette smoking, and education were adjusted. OR (95% CI) for age indicates the risk of outcome per 10-year age increase. OR=odds ratio. 95% CI=95% confidence interval.

Among 67 patients with two measurements, 38 (57%) had sustained PNP twice; 11(16%) had no PNP in 6-month measurement but developed PNP in 12-month measurement; 8(12%) never had PNP; 10 (15%) had PNP in 6-month measurement and no PNP in 12-month measurement. Result of univariable logistic regression showed age was associated with increased risk of sustained or developing PNP in 12-month follow-up (OR=1.38 per 10-year increase of age, 95% CI, 1.03-1.85), while no significant association in multivariable analysis (OR=1.30, 95% CI, 0.96-1.77, eTable 7).

As shown in Table 2, the amplitudes of the motor branch of the ulnar nerve were all significantly increased, and the left wrist segment was 4.9 ± 1.1 mv for 6-month measurement vs 5.3 ± 1.2 mv for 12-month measurement; left elbow segment was 4.7 ± 1.1 mv vs 5.2 ± 1.2 mv; right wrist segment was 4.8 ± 1.3 mv vs 5.4 ± 1.4 mv; right elbow segment was 4.7 ± 1.4 mv vs 5.4 ± 1.4 mv. Sural nerve amplitude also significantly increased, with left side 3.6 ± 1.2 mv vs 4.2 ± 1.5 mv, and right side 3.7 ± 1.3 mv vs 4.3 ± 1.8 mv. The amplitude of the motor branch of the median nerve in the left wrist also increased, which was 6.1 ± 1.6 mv and 6.4 ± 1.8mv. No significant difference of amplitude in peroneal nerve or tibial nerve between two measurements was observed (Table 2). The conduction velocity of the right carpal motor branch of the median nerve decreased and was 54.2 ± 10.3 m/s vs 50.2 ± 11.1 m/s, respectively. The velocity of the sensory branch of the right median nerve also decreased and was 51.2 ± 9.4 m /s vs 49.1 ± 8.6 m/s respectively. The CV of peroneal nerve in the left fibular head segment increased and was 49.6 ± 6.9 m/s vs. 53.1 ± 6.4 m/s respectively.

**Table 2:**
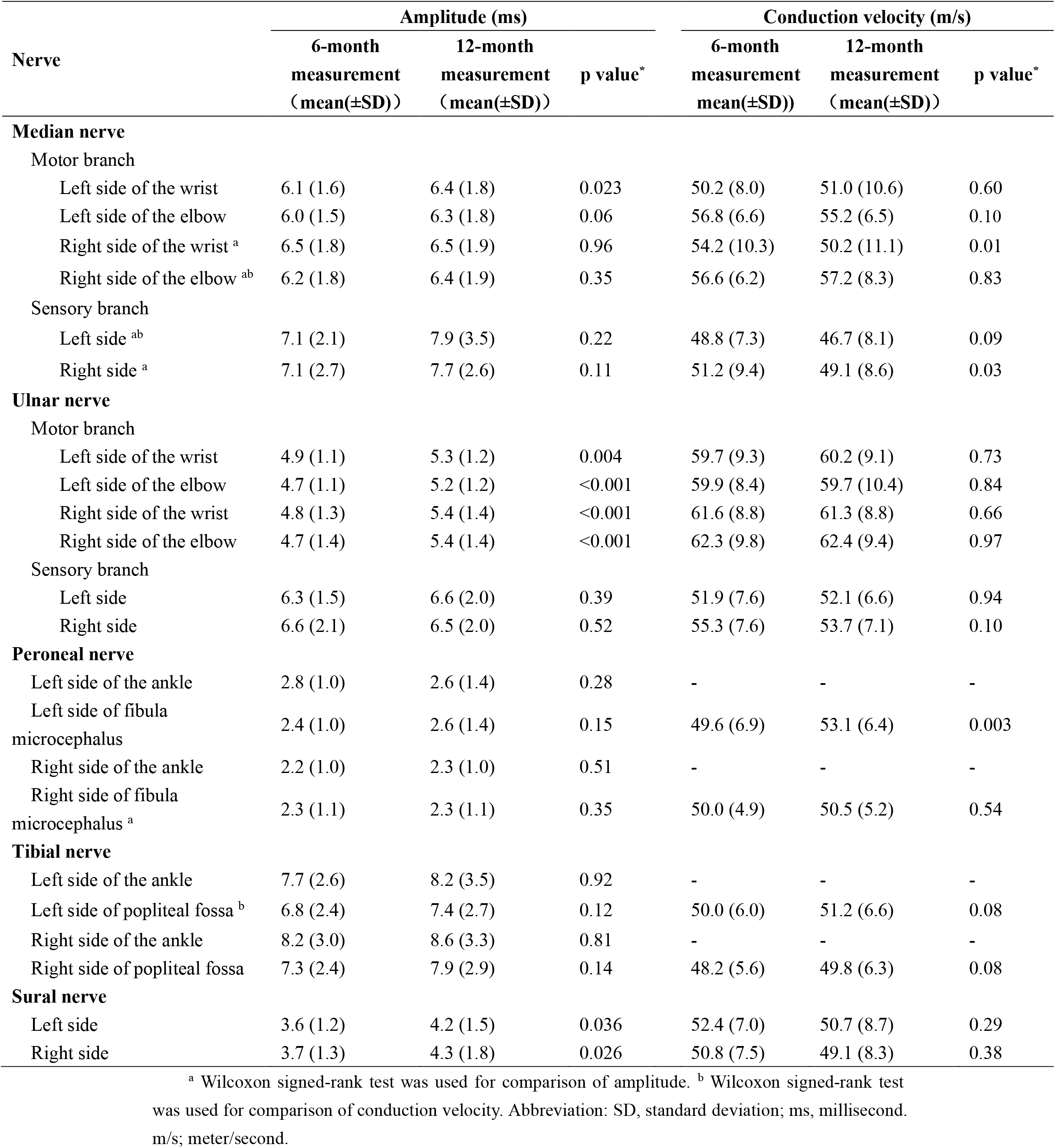
Comparison of amplitude and conduction velocity of two measurements among 67 patients.

## Discussion

In this study, we found in hospital survivors with COVID-19, the main accompanying symptoms were memory loss, hair loss, anxiety, sleep difficulties, and fatigue. Most patients (74%) had PNP, including 51 patients (16%) with mononeuropathy and 181 patients (58%) with generalized PNP. Patients at 12-month measurement had a higher prevalence of generalized PNP. 64 (20%) patients had only axonal loss, 67 (21%) had only demyelination, and 101 (32%) had a mixed type. Additionally, we found that the main influencing factors were age, education, and antibody IgG level at discharge.

### Research Status of hospital survivors with COVID-19 and effects on Nervous System

Huang et al.^12^ found that among 2,469 COVID-19 patients, most of them had at least one sequelae symptom at 6 months after discharge and its result was consistent with our findings. In our study, we found no significant association between any symptoms and PNP, which suggest all the COVID-19 survivors should pay attention to the observation of peripheral nerves during follow-up.

We found that most patients were found with PNP, which was consistent with previous studies. Oaklander^14^ found among 17 long-term COVID-19 patients, 59% of them had ≥1 test which confirmed the presence of PNP. Guerrero et al^15^ reviewed and summarized 143 studies about nervous system involvement by COVID-19. A total of 10,723 COVID-19 patients were reported to be affected by SARS-CoV-2 in the central and peripheral nervous systems. The study reflected the remarkable prevalence of neurological involvement in COVID-19 patients and the rate of neurological involvement ranged from 22.5% to 36.4%. However, previous studies on the nervous system mainly focused on central nervous system ^16-18^, and most of the studies on PNP were case reports or focused on severe COVID-19 patients^19^.

In the studies mentioned above, electrophysiological testing was not performed, which may be related to the insufficient use of the equipment for electrophysiological examination. Additionally, PNP is relatively insidious, the clinical manifestations are not typical, and many cases gradually appear after acute infection. These reasons have resulted in the lack of large-scale studies of PNP in COVID-19 patients, especially on neurological recovery in long-term rehabilitation of the COVID-19 survivors.

NCS is not only important for the diagnosis of peripheral neuropathy, but also for prognosis and selection of the best therapy^20^. A previous study^21^ assessed the correlation between electromyography (EMG) and NCS in COVID-19 patients and found that NCS was important for predicting COVID-19 prognosis and recovery.

### Characteristics and Influencing Factors of NCS in hospital survivors with COVID-19

By using NCS, we found that most of the patients had persistent PNP, mainly for generalized PNP at 6 or 12 months after infection with SARS-CoV-2. PNP was positively associated with age. Prevalence of F-wave abnormality was relatively low and patients with high levels of IgG antibody at discharge had a reduced risk of F-wave abnormality. Several studies^22-24^ have confirmed that IgG antibodies play an important role in neutralizing the virus and protecting the body from virus reinfection. Therefore, timely COVID-19 vaccination and booster vaccination to improve the antibody level may help reduce the prevalence and severity of PNP.

Through two measurements, we found among the pathological types, the mixed type (axonal loss combined with demyelination) had the highest rate. The prevalence of median nerve injured was highest and both the motor and sensory branches were damaged, with demyelination mainly, and distal (carpal-palm) involvement; followed by the peroneal nerve with mainly axonal loss and in both proximal and distal ends. Compared with the measurement at 6-month follow-up, amplitude of the ulnar nerve and sural nerve increased, while the conduction velocity of the median nerve was decreased of 67 patients who completed 2 measurements, which indicated axonal loss of the ulnar nerve and sural nerve was relieved, and the demyelination of the median nerve was aggravated.

The results above suggest that COVID-19 patients are more prone to have PNP in the distal limb and had a higher prevalence in lower extremities especially in the long term of rehabilitation. After the diagnosis of COVID-19 infection, we should pay more attention to the peripheral nerve condition of patients with old age and low IgG antibody level, especially those bedridden and immobilized. Additionally, it reminds us to strengthen COVID-19 vaccination to avoid more severe neurological damage.

### Possible Pathogenesis of PNP in COVID-19 survivors

However, the specific mechanism is still unclear, and the current research focuses on the following three aspects:

#### Direct Neuroinvasive Effects of SARS-CoV-2

The nervous system damage of COVID-19 patients is mostly caused by acute nervous system infection due to SARS-CoV-2 virus^25^. Infection with SARS-CoV-2 puts patients at increased risk of neurological damage, and in some cases SARS-CoV-2 causes clinical manifestations of acute neurological damage or exacerbates pre-existing baseline neurological disease severity. Another factor to consider is the possible direct neuro-muscular damage caused by the SARS-CoV-2 virus. However, direct muscle damage by the virus has not been demonstrated, nor has it been reported in autologous muscle lesions.

#### Autoimmune Response

Finsterer J^26^ analyzed 105 studies on SARS-CoV-2-related PNP. It was speculated that the infected virus may have epitopes similar to components of the peripheral nerve (activating autoreactive B or T cells) and result in PNP. SARS-CoV-2 spikes interact with GM1 gangliosides in peripheral nerves, leading to cross-reactivity and production of antibodies against these antigens, inducing a peripheral demyelinating pattern of GBS^27^.

#### Persistent and Recurring Neuroinflammatory Responses and Damage

After infection with SARS-CoV-2, immune dysregulation may persist in the form of persistent inflammation, immunosuppression and catabolic syndrome^28^. This immune state is thought to be triggered by cytokine storms during acute infection and further contributed by the sustained release of endogenous alarmins or danger-associated molecular patterns, which can lead to chronic systemic inflammation. Chronic neuroinflammation associated with high levels of cytokines/chemokines may be involved in the pathogenesis and progression of neurological diseases^27^.

### Limitations and Prospects

This study is only a preliminary analysis of the peripheral nerve

electrophysiology-related indicators and baseline data of patients in the recovery period of patients with COVID-19, trying to find the characteristics and related risk factors of PNP among COVID-19 survivors in the medium- and long-term rehabilitation. This study does not describe the intrinsic link between persistent multi-organ functional injury and PNP. In the ongoing second part of the study, we will also comprehensively analyze more clinical data of COVID-19 patients during hospitalization and discharge, i.e., the blood biochemical test indicators, lung function, six-minute walk test, etc. Correlation analysis with the relevant indicators of peripheral nerve electrophysiology will be carried out to deeply reveal the relationship between PNP and the function of major organs of the body and provide real data for the complete recovery.

## Data Availability

All data produced in the present study are available upon reasonable request to the authors

## Author Contributions

Dr Zhao had full access to all the data in the study and takes responsibility for the integrity of the data and the accuracy of the data analysis. Qian Yang and Meng Zhang contributed equally. Drs Zhao and Sun contributed equally.

Study design: Zhao, Sun.

Statistical analysis: Zhao, Sun, Yang, Zhang.

Acquisition of data: Zhao, Yang, Lai, X Liu, F Liu, Li, Zang, Song, Ma, Wang, Cui.

Critical revision of the manuscript for important intellectual content: All authors.

Drafting of the manuscript: Zhao, Sun, Yang, Zhang, Shao.

