## Supplemental Table 1-7 for "COVID-19 could cause long term peripheral nerve demyelination and axonal loss: A One Year Prospective Cohort Study"

**People's Hospital, Shijiazhuang, Hebei Province, China** (Yang, Lai, Li, Cui, Ma, Wang, Cui, Zhao); **Department of Epidemiology and Health Statistics, Peking University, Beijing, China** (Zhang, Shao, Sun); **North China University of Science and Technology, Tangshan, Hebei Province, China** (X Liu); **Hebei University of Chinese Medicine, Shijiazhuang, Hebei Province, China** (F Liu, Zang, Song)

**Correspondence to:**

Yubin Zhao, People's Hospital, Shijiazhuang, Hebei Province 050000, China

or

Feng Sun, Department of Epidemiology and Health Statistics, Peking University, Beijing 100191, China

Word count: 3206

**Abstract**

**Background:** There is a lack of studies on large-sample, medium-, or long-term follow-up data of peripheral neuropathy (PNP) in the COVID-19 survivors. This study evaluated the characteristics and related risk factors of PNP in the medium- and long-term rehabilitation,which provided real-world study data for the complete recovery of COVID-19 patients.

**Methods:** This study was a prospective cohort study of the COVID-19 survivors. We collected data on baseline characteristics, symptoms at onset and after discharge during the 6-month and 12-month follow-up. Peripheral nerves were measured by electromyography and inducible potentiometer. We used multivariable logistic regression to analyze the influencing factors of PNP. Additionally, we compared the difference between the two measurements among the population who completed both measurements.

**Results:** 313 patients were included in the study and all of them underwent nerve conduction study. 67 patients completed two measurements at 6-month and 12-month follow-up. Commonly reported symptoms contained memory loss (86%), hair loss (28%), anxiety (24%), and sleep difficulties (24%). 232 patients (74%) were found with PNP, including 51 (16%) with mononeuropathy and 181 (58%) with generalized PNP. Patients with measurement at 12-month follow-up had a higher prevalence of generalized PNP (p=0.006). For pathological types, 64 (20%) patients had only axonal loss, 67 (21%) had only demyelination, and 101 (32%) had a mixed type. There was no significant difference in the prevalence of accompanying symptoms after discharge between the two groups with or without PNP. After adjustment, age was positively associated with PNP (OR=1.22 per 10-year increase of age, 95% CI, 1.05-1.41). Compared with less than the median amount of IgG at discharge, higher amount of IgG was associated with decreased risk of F-wave abnormality (OR=0.32, 95%CI, 0.11-0.82), but no significant difference in other types of PNP.

**Conclusions and Relevance:** SARS-CoV-2 could cause PNP in hospital survivors with COVID-19, which persisted and was associated with age, education, and IgG antibody at discharge, but had no significant correlation with symptoms after discharge.

### Supplemental Online Content

**eMethods.** Procedures and criteria of nerve conduction study

**eTable 1.** Baseline characteristics of 67 patients with two measurements

**eTable 2.** Prevalence and characteristics of peripheral neuropathy

**eTable 3.** Prevalence of peripheral neuropathy in two or more types of nerve injured

**eTable 4.** Pathological classification of peripheral neuropathy of each nerve

**eTable 5.** Prevalence of symptoms after discharge in COVID-19 patients

**eTable 6.** Risk factors of peripheral neuropathy in univariable logistic analysis

**eTable 7.** Risk factors of peripheral neuropathy in logistic analysis among 67 patients with two measurements

**eMethods**

**Procedures and criteria of nerve conduction study**

(1) Skin temperature: before testing, ensure that the skin temperature is between 30 and 32°C. (2) Electrodes: we used disk-shaped surface electrodes and recorded the compound muscle action potential (CMAP) and sensory nerve action potential (SNAP). (3) Electrode placement: i Stimulate electrode: when measuring motor conduction, the cathode was placed at the distal end, and the anode was placed at the proximal end; and when F-wave was measured, the cathode was placed at the proximal end. In retrograde sensory conduction measurement, the stimulating electrode was placed on the nerve stem, the cathode was at the distal end, and the anode was at the proximal end. The distance between the cathode and the anode was about 2 cm; ii Record electrode: when motor conduction was measured, the acting electrode was placed on the belly of the muscle, and the reference electrode was placed on the tendon or its attachment point near the muscle. iii Ground wire: it was placed between the stimulating electrode and the recording electrode. (4) Stimulation intensity and time limit: during motor conduction measurement, super-stimulation was applied to the nerve stem to increase the stimulation intensity by 10% to 30% to induce the maximum CMAP stimulation intensity. Stimulus duration was 0.1ms or 0.2ms.

The measurement method of the F-wave was similar with that of the motor conduction measurement, while the cathode of the stimulating electrode is placed at the proximal end.

#### eTable 1: Baseline characteristics of 67 patients with two measurements

| Characteristics | Total (n=67) | 6-month (n=67) | | | 12-month (n=67) | | |
|---|---|---|---|---|---|---|---|
| | | No peripheral neuropathy (n=19) | Peripheral neuropathy (n=48) | p value | No peripheral neuropathy (n=18) | Peripheral neuropathy (n=49) | p value |
| Age, mean ± SD, years | 45.6 ± 18.4 | 43.2 ± 18.2 | 46.6 ± 18.6 | 0.49 | 37.6 ± 17.9 | 48.6 ± 17.8 | 0.03 |
| Age, median (IQR), years | 50.0 (34.5-59.0) | 48.0 (33.0-57.5) | 50.5 (38.3-62.0) | 0.44 | 38.5 (31.3-51.8) | 53.0 (41.0-62.0) | 0.01 |
| Sex | | | | 0.94 | | | 0.39 |
| Male | 26 (39%) | 8 (42%) | 18 (38%) | | 9 (50%) | 17 (35%) | |
| Female | 41 (61%) | 11 (58%) | 30 (63%) | | 9 (50%) | 32 (65%) | |
| Smoking | | | | 0.85 | | | 0.58 |
| Never-smoker | 59 (88%) | 16 (84%) | 43 (90%) | | 17 (94%) | 42 (86%) | |
| Current or former smoker | 8 (12%) | 3 (16%) | 5 (10%) | | 1 (6%) | 7 (14%) | |
| Education | | | | 0.14 | | | 1.00 |
| Middle school or lower | 65 (97%) | 17 (90%) | 48 (100%) | | 17 (94%) | 48 (98%) | |
| College or higher | 2 (3%) | 2 (11%) | 0 (0%) | | 1 (6%) | 1 (2%) | |
| Disease severity | | | | 0.52 | | | 0.08 |
| Mild | 16 (24%) | 3 (16%) | 13 (27%) | | 2 (11%) | 14 (29%) | |
| Moderate | 46 (69%) | 15 (79%) | 31 (65%) | | 16 (89%) | 30 (61%) | |
| Severe or critically ill | 5 (8%) | 1 (5%) | 4 (8%) | | 0 (0%) | 5 (10%) | |
| Symptoms and signs at disease onset | | | | | | | |
| Fever | 11 (16%) | 2 (11%) | 9 (19%) | 0.65 | 4 (22%) | 7 (14%) | 0.69 |
| Cough | 16 (24%) | 4 (21%) | 12 (25%) | 0.98 | 4 (22%) | 12 (25%) | 0.99 |
| Dyspnea | 3 (5%) | 0 (0%) | 3 (6%) | 0.65 | 1 (6%) | 2 (4%) | 0.99 |
| Fatigue | 3 (5%) | 2 (11) | 1 (2%) | 0.40 | 0 (0%) | 3 (6%) | 0.68 |
| Sputum production | 4 (6%) | 1 (5%) | 3 (6%) | 0.99 | 2 (11%) | 2 (4%) | 0.62 |
| Hypertension | 13 (19%) | 1 (5%) | 12 (25%) | 0.13 | 2 (11%) | 11 (22%) | 0.49 |
| Antiviral therapy | 5 (8%) | 1 (5%) | 4 (8%) | 0.99 | 2 (11%) | 3 (6%) | 0.87 |
| nCoV IgG at discharge, S/CO | 31.0 (15.4-120.7) | 26.9 (13.3-191.8) | 32.2 (18.0-111.9) | 0.93 | 25.9 (13.7-125.2) | 32.3 (18.5-114.2) | 0.70 |
| Time from admission to 6-month follow-up, median (IQR), days | 151.0 (147.5-154.0) | 152.0 (148.5-153.5) | 151.0 (147.0-154.0) | 0.45 | 151.5 (149.3-154.8) | 151.0 (147.0-154.0) | 0.20 |
| Time from admission to 12-month follow-up, median (IQR), days | 352.0 (346.0-356.0) | 352.0 (345.5-355.5) | 353.0 (346.0-356.3) | 0.64 | 354.0 (350.5-356.0) | 352.0 (346.0-356.0) | 0.35 |
| Length of hospital stay, median (IQR), days | 17.0 (15.0-24.0) | 21.0 (15.5-26.0) | 17.0 (14.0-23.0) | 0.19 | 18.0 (15.0-22.5) | 17.0 (14.0-24.0) | 0.92 |

SD=standard deviation. IQR=interquartile range. nCoV IgG=novel coronavirus immunoglobulin G. S/CO=sample/cut off.

#### eTable 2: Prevalence and characteristics of peripheral neuropathy

| | Total （n=313） | 6-month follow-up (n=83) | 12-month follow-up (n=230) | p value |
|---|---|---|---|---|
| **Peripheral neuropathy** | | | | 0.08 |
| Yes | 232 (74%) | 55 (66%) | 177 (77%) | |
| No | 81 (26%) | 28 (34%) | 53 (23%) | |
| **Classification** | | | | |
| Mononeuropathy | 51 (16%) | 18 (22%) | 33 (14%) | 0.17 |
| Polyneuropathy | 181 (58%) | 37 (45%) | 144 (63%) | 0.006 |
| **Nerve Injured** | | | | |
| **Median nerve** | 135 (43%) | 31 (37%) | 104 (45%) | 0.27 |
| Different branch | | | | 0.55 |
| Motor branch only | 34 (11%) | 7 (8%) | 27 (12%) | |
| Sensory branch only | 31 (10%) | 6 (7%) | 25 (11) | |
| Motor and Sensory | 70 (22%) | 18 (22%) | 52 (23%) | |
| **Ulnar nerve** | 64 (20%) | 19 (23%) | 45 (20%) | 0.63 |
| Different branch | | | | 0.69 |
| Motor branch only | 45 (14%) | 15 (18%) | 30 (13%) | |
| Sensory branch only | 14 (5%) | 3 (4%) | 11 (5%) | |
| Motor and Sensory | 5 (2%) | 1 (1%) | 4 (2%) | |
| **Peroneal nerve** | 111 (36%) | 24 (29%) | 87 (38%) | 0.19 |
| **Tibial nerve** | 77 (25%) | 12 (15%) | 65 (28%) | 0.02 |
| **Sural nerve** | 56 (18%) | 9 (11%) | 47 (20%) | 0.07 |
| **Nerve injured in different areas** | | | | 0.12 |
| Nerve injured in one area | 51 (16%) | 18 (22%) | 33 (14%) | |
| Nerve injured in two areas | 73 (23%) | 18 (22%) | 55 (24%) | |
| Nerve injured in three areas | 34 (11%) | 3 (4%) | 31 (14%) | |
| Nerve injured in four areas | 34 (11%) | 8 (10%) | 26 (11%) | |
| Nerve injured in five areas | 22 (7%) | 5 (6%) | 17 (7%) | |
| Nerve injured in six areas | 11 (4%) | 1 (1%) | 10 (4%) | |
| Nerve injured in seven areas | 6 (2%) | 2 (2%) | 4 (2%) | |
| Nerve injured in eight areas | 1 (0%) | 0 (0%) | 1 (0%) | |
| **Number of different types of nerve injured*** | | | | 0.09 |
| One type of nerve injured | 95 (30%) | 30 (36%) | 65 (28%) | |
| Two types of nerves injured | 81 (26%) | 15 (18%) | 66 (29%) | |
| Three types of nerves injured | 41 (13%) | 6 (7%) | 35 (15%) | |
| Four types of nerves injured | 12 (4%) | 3 (4%) | 9 (4%) | |
| Five types of nerves injured | 3 (1%) | 1 (1%) | 2 (1%) | |
| **Pathological classification** | | | | 0.17 |
| Demyelination only | 67 (21%) | 18 (22%) | 49 (21%) | |
| Axonal loss only | 64 (20%) | 17 (21%) | 47 (20%) | |
| Demyelination combined with axonal loss | 101 (32%) | 20 (24%) | 81 (35%) | |
| **Peripheral neuropathy on different side of body** | | | | 0.24 |
| Left side only | 32 (10%) | 7 (8%) | 25 (11%) | |
| Right side only | 23 (7%) | 7 (8%) | 16 (7%) | |
| Both sides | 177 (57%) | 41 (49%) | 136 (59%) | |

| | Total（n=313） | 6-month follow-up (n=83) | 12-month follow-up (n=230) | p value |
|---|---|---|---|---|
| **Classification by anatomical location of motor branch** | | | | |
| Proximal limb only | 0 (0%) | 0 (0%) | 0 (0%) | - |
| Distal limb only | 53 (17%) | 15 (18%) | 38 (17%) | 0.88 |
| Proximal and distal limbs | 164 (52%) | 38 (46%) | 126 (55%) | 0.20 |
| **F-wave abnormality** | 22 (7%) | 3 (4%) | 19 (8%) | 0.24 |
| **F-wave abnormality on different side of body** | | | | 0.41 |
| Left side only | 12 (4%) | 1 (1%) | 11 (5%) | |
| Right side only | 8 (3%) | 2 (2%) | 6 (3%) | |
| Both sides | 2 (1%) | 0 (0%) | 2 (1%) | |
| **Nerve with F-wave abnormality** | | | | 0.50 |
| Median nerve only | 19 (6%) | 3 (4%) | 16 (7%) | |
| Ulnar nerve only | 2 (1%) | 0 (0%) | 2 (1%) | |
| Median and ulnar nerve | 1 (0%) | 0 (0%) | 1 (0%) | |

* One type of nerve contains the same nerve on both sides of body.

#### eTable 3: Prevalence of peripheral neuropathy in two or more types of nerve injured

| | Total (n=313) | 6-month follow-up (n=83) | 12-month follow-up (n=230) | p value |
|---|---|---|---|---|
| **Two nerves injured at the same time** | 81 (26%) | 15 (18%) | 66 (29%) | 0.08 |
| Median nerve & Ulnar nerve | 10 (3%) | 2 (2%) | 8 (4%) | 0.91 |
| Median nerve & Peroneal nerve | 21 (7%) | 6 (7%) | 15 (7%) | 0.99 |
| Median nerve &Tibial nerve | 9 (3%) | 1 (1%) | 8 (4%) | 0.50 |
| Median nerve & Sural nerve | 11 (4%) | 1 (1%) | 10 (4%) | 0.32 |
| Ulnar nerve & Peroneal nerve | 5 (2%) | 1 (1%) | 4 (2%) | 0.99 |
| Ulnar nerve & Tibial nerve | 4 (1%) | 0 (0%) | 4 (2%) | 0.52 |
| Ulnar nerve & Sural nerve | 4 (1%) | 1 (1%) | 3 (1%) | 0.99 |
| Peroneal nerve & Tibial nerve | 11 (4%) | 2 (2%) | 9 (4%) | 0.77 |
| Peroneal nerve & Sural nerve | 5 (2%) | 0 (0%) | 5 (2%) | 0.40 |
| Tibial nerve & Sural nerve | 1 (0%) | 1 (1%) | 0 (0%) | 0.59 |
| **Three nerves injured at the same time** | 41 (13%) | 6 (7%) | 35 (15%) | 0.10 |
| Median nerve & Ulnar nerve & Peroneal nerve | 5 (2%) | 1 (1%) | 4 (2%) | 0.99 |
| Median nerve & Ulnar nerve & Tibial nerve | 3 (1%) | 1 (1%) | 2 (1%) | 0.99 |
| Median nerve & Ulnar nerve & Sural nerve | 2 (1%) | 1 (1%) | 1 (0%) | 0.99 |
| Median nerve & Peroneal nerve & Tibial nerve | 11 (4%) | 0 (0%) | 11 (5%) | 0.09 |
| Median nerve & Peroneal nerve & Sural nerve | 8 (3%) | 1 (1%) | 7 (3%) | 0.61 |
| Median nerve &Tibial nerve& Sural nerve | 4 (1%) | 0 (0%) | 4 (2%) | 0.52 |
| Ulnar nerve & Peroneal nerve & Tibial nerve | 4 (1%) | 1 (1%) | 3 (1%) | 0.99 |
| Ulnar nerve & Peroneal nerve & Sural nerve | 1 (0%) | 1 (1%) | 0 (0%) | 0.59 |
| Ulnar nerve & Tibial nerve & Sural nerve | 1 (0%) | 0 (0%) | 1 (0%) | 0.99 |
| Peroneal nerve & Tibial nerve & Sural nerve | 2 (1%) | 0 (0%) | 2 (1%) | 0.96 |
| **Four nerves injured at the same time** | 12 (4%) | 3 (4%) | 9 (4%) | 0.99 |
| Five nerves except for the median nerve | 0 (0%) | 0 (0%) | 0 (0%) | - |
| Five nerves except for the Tibial nerve | 4 (1%) | 1 (1%) | 3 (1%) | 0.99 |
| Five nerves except for the Peroneal nerve | 1 (0%) | 0 (0%) | 1 (0%) | 0.99 |
| Five nerves except for the Tibial nerve | 3 (1%) | 1 (1%) | 2 (1%) | 0.99 |
| Five nerves except for the Sural nerve | 4 (1%) | 1 (1%) | 3 (1%) | 0.99 |
| **Five nerves injured at the same time** | 3 (1%) | 1 (1%) | 2 (1%) | 0.99 |

**eTable 4: Pathological classification of peripheral neuropathy of each nerve**

| Pathological classification | Total （n=313） | 6-month follow-up (n=83) | 12-month follow-up (n=230) | p value |
|---|---|---|---|---|
| **Median nerve** | | | | 0.40 |
| Demyelination only | 130 (42%) | 31 (37%) | 99 (43%) | |
| Axonal loss only | 2 (1%) | 0 (0%) | 2 (1%) | |
| Demyelination combined with axonal loss | 3 (1%) | 0 (0%) | 3 (1%) | |
| **Ulnar nerve** | | | | 0.44 |
| Demyelination only | 14 (5%) | 3 (4%) | 11 (5%) | |
| Axonal loss only | 47 (15%) | 16 (19%) | 31 (14%) | |
| Demyelination combined with axonal loss | 3 (1%) | 0 (0%) | 3 (1%) | |
| **Peroneal nerve** | | | | 0.02 |
| Demyelination only | 8 (3%) | 5 (6%) | 3 (1%) | |
| Axonal loss only | 96 (31%) | 17 (21%) | 79 (34%) | |
| Demyelination combined with axonal loss | 7 (2%) | 2 (2%) | 5 (2%) | |
| **Tibial nerve** | | | | 0.06 |
| Demyelination only | 16 (5%) | 2 (2%) | 14 (6%) | |
| Axonal loss only | 55 (18%) | 8 (10%) | 47 (20%) | |
| Demyelination combined with axonal loss | 6 (2%) | 2 (2%) | 4 (2%) | |
| **Sural nerve*** | | | | 0.16 |
| Demyelination only | 53 (17%) | 8 (10%) | 45 (20%) | |
| Axonal loss only | 1 (0%) | 0 (0%) | 1 (0%) | |
| Demyelination combined with axonal loss | 1 (0%) | 0 (0%) | 1 (0%) | |

*Both side of sural nerves were not detected in one patient.

#### eTable 5: Prevalence of symptoms after discharge in COVID-19 patients

| Symptoms | Total (n=313) | No peripheral neuropathy (n=81) | Peripheral neuropathy (n=232) | p value |
|---|---|---|---|---|
| Fever | 2 (1%) | 1 (1%) | 1 (0%) | 0.99 |
| Cough | 55 (18%) | 16 (20%) | 39 (17%) | 0.67 |
| Expectoration | 45 (14%) | 12 (15%) | 33 (14%) | 0.99 |
| Dry throat | 65 (21%) | 20 (25%) | 45 (19%) | 0.39 |
| Pharyngeal itching | 36 (12%) | 11 (14%) | 25 (11%) | 0.63 |
| Congestion in the throat | 33 (11%) | 9 (11%) | 24 (10%) | 0.99 |
| Sore throat | 14 (5%) | 6 (7%) | 8 (3%) | 0.24 |
| Heterorexia | 9 (3%) | 2 (3%) | 7 (3%) | 0.99 |
| Olfactory abnormality | 24 (8%) | 9 (11%) | 15 (7%) | 0.27 |
| Muscle weakness | 66 (21%) | 14 (17%) | 52 (22%) | 0.41 |
| Hyperhidrosis | 51 (16%) | 14 (17%) | 37 (16%) | 0.92 |
| Aversion to wind | 52 (17%) | 13 (16%) | 39 (17%) | 0.99 |
| Shortness of breath | 56 (18%) | 14 (17%) | 42 (18%) | 0.99 |
| Chest tightness | 47 (15%) | 11 (14%) | 36 (16%) | 0.81 |
| Chest pain | 12 (4%) | 3 (4%) | 9 (4%) | 0.99 |
| Physical pain | 37 (12%) | 11 (14%) | 26 (11%) | 0.71 |
| Palpitation | 45 (14%) | 10 (12%) | 35 (15%) | 0.67 |
| Dysphoria | 75 (24%) | 19 (24%) | 56 (24%) | 0.99 |
| Sleep disorders | 74 (24%) | 18 (22%) | 56 (24%) | 0.84 |
| Hair loss | 86 (28%) | 23 (28%) | 63 (27%) | 0.94 |
| Dizziness | 41 (13%) | 10 (12%) | 31 (13%) | 0.97 |
| Headache | 29 (9%) | 10 (12%) | 19 (8%) | 0.37 |
| Weakness of lower limbs | 37 (12%) | 8 (10%) | 29 (13%) | 0.67 |
| Bitter taste | 65 (21%) | 14 (17%) | 51 (22%) | 0.46 |
| Nausea | 12 (4%) | 4 (5%) | 8 (3%) | 0.79 |
| Decreased appetite | 12 (4%) | 2 (3%) | 10 (4%) | 0.68 |
| Constipation | 38 (12%) | 5 (6%) | 33 (14%) | 0.09 |
| Diarrhea | 21 (7%) | 7 (9%) | 14 (6%) | 0.58 |
| Abnormal urination | 23 (7%) | 4 (5%) | 19 (8%) | 0.47 |
| Fatigue | 63 (20%) | 13 (16%) | 50 (22%) | 0.37 |
| Memory Loss | 269 (86%) | 74 (91%) | 195 (84%) | 0.15 |

#### eTable 6: Risk factors of peripheral neuropathy in univariable logistic analysis

| | Peripheral neuropathy | | Mononeuropathy | | Generalized peripheral neuropathy | | F-wave abnormality | |
|---|---|---|---|---|---|---|---|---|
| | OR (95%CI) | p value | OR (95%CI) | p value | OR (95%CI) | p value | OR (95%CI) | p value |
| **Age*** | 1.24 (1.07-1.43) | 0.004 | 1.00 (0.84-1.19) | 1.00 | 1.19 (1.04-1.35) | 0.010 | 1.00 (0.78-1.29) | 0.98 |
| **Sex** | | | | | | | | |
| Men | 1 (ref) | | 1 (ref) | | 1 (ref) | | 1 (ref) | |
| Women | 1.56 (0.93-2.60) | 0.09 | 1.09 (0.59-2.06) | 0.78 | 1.36 (0.86-2.15) | 0.19 | 1.40 (0.57-3.76) | 0.48 |
| **Education** | | | | | | | | |
| Middle school or lower | 1 (ref) | | 1 (ref) | | 1 (ref) | | 1 (ref) | |
| College or higher | 0.74 (0.28-2.18) | 0.56 | 4.25 (1.56-11.10) | 0.003 | 0.24 (0.08-0.64) | 0.008 | 0.72 (0.04-3.78) | 0.76 |
| **Smoking** | | | | | | | | |
| Never-smoker | 1 (ref) | | 1 (ref) | | 1 (ref) | | 1 (ref) | |
| Current or former smoker | 1.16 (0.50-3.03) | 0.74 | 1.03 (0.33-2.63) | 0.95 | 1.10 (0.52-2.44) | 0.80 | 0.43 (0.02-2.18) | 0.42 |
| **Comorbidity** | | | | | | | | |
| No | 1 (ref) | | 1 (ref) | | 1 (ref) | | 1 (ref) | |
| Yes | 1.34 (0.74-2.53) | 0.34 | 0.80 (0.37-1.60) | 0.55 | 1.42 (0.84-2.43) | 0.20 | 1.14 (0.40-2.89) | 0.79 |
| **nCoV IgG at discharge** | | | | | | | | |
| < median | 1 (ref) | | 1 (ref) | | 1 (ref) | | 1 (ref) | |
| ≥ median | 1.46 (0.88-2.44) | 0.15 | 1.38 (0.76-2.55) | 0.30 | 1.12 (0.72-1.76) | 0.61 | 0.35 (0.12-0.87) | 0.03 |
| **Antiviral** | | | | | | | | |
| No | 1 (ref) | | 1 (ref) | | 1 (ref) | | 1 (ref) | |
| Yes | 2.00 (0.80-6.07) | 0.17 | 1.51 (0.57-3.54) | 0.37 | 1.24 (0.59-2.71) | 0.57 | 0.40 (0.02-2.02) | 0.38 |

| | Peripheral neuropathy | | Mononeuropathy | | Generalized peripheral neuropathy | | F-wave abnormality | |
|---|---|---|---|---|---|---|---|---|
| | **OR (95%CI)** | **p value** | **OR (95%CI)** | **p value** | **OR (95%CI)** | **p value** | **OR (95%CI)** | **p value** |
| **Corticosteriods** | | | | | | | | |
| No | 1 (ref) | | 1 (ref) | | 1 (ref) | | 1 (ref) | |
| Yes | 0.87 (0.18-6.16) | 0.87 | 0.85 (0.04-5.14) | 0.88 | 0.97 (0.21-5.00) | 0.97 | 2.26 (0.12-14.11) | 0.46 |
| **Length of hospital stay** | | | | | | | | |
| ≤14 days | 1 (ref) | | 1 (ref) | | 1 (ref) | | 1 (ref) | |
| 15-21 days | 0.90 (0.49-1.65) | 0.75 | 0.95 (0.45-2.03) | 0.89 | 0.95 (0.55-1.63) | 0.85 | 1.67 (0.59-5.40) | 0.35 |
| >21 days | 1.16 (0.59-2.31) | 0.67 | 1.23 (0.57-2.71) | 0.60 | 0.99 (0.55-1.80) | 0.98 | 0.98 (0.26-3.63) | 0.97 |

*OR (95% CI) for age indicates the risk of outcome per 10-year age increase. OR=odds ratio. 95%CI=95% confidence interval.

#### eTable 7: Risk factors of peripheral neuropathy in logistic analysis among 67 patients with two measurements

| | Univariable | | Multivariable | |
|---|---|---|---|---|
| | OR (95%CI) | p value | OR (95%CI) | p value |
| **Age*** | 1.38 (1.03-1.85) | 0.03 | 1.30 (0.96-1.77) | 0.09 |
| **Sex** | | | | |
| Men | 1 (ref) | | 1 (ref) | |
| Women | 1.88 (0.63-5.72) | 0.26 | 1.75 (0.39-7.78) | 0.45 |
| **Education** | | | | |
| Middle school or lower | 1 (ref) | | 1 (ref) | |
| College or higher | 0.35 (0.01-9.29) | 0.47 | 0.38 (0.01-10.68) | 0.52 |
| **Smoking** | | | | |
| Never-smoker | 1 (ref) | | 1 (ref) | |
| Current or former smoker | 2.83 (0.45-55.06) | 0.35 | 2.57 (0.31-56.23) | 0.44 |
| **Comorbidity** | | | | |
| No | 1 (ref) | | 1 (ref) | |
| Yes | 2.32 (0.54-16.07) | 0.31 | 1.68 (0.28-14.73) | 0.60 |
| **nCoV IgG at discharge** | | | | |
| < median | 1 (ref) | | 1 (ref) | |
| ≥ median | 1.41 (0.48-4.29) | 0.53 | 1.62 (0.49-5.77) | 0.44 |
| **Antiviral** | | | | |
| No | 1 (ref) | | 1 (ref) | |
| Yes | 0.52 (0.08-4.23) | 0.50 | 0.51 (0.05-5.61) | 0.56 |
| **Length of hospital stay** | | | | |
| ≤14 days | 1 (ref) | | 1 (ref) | |
| 15-21 days | 0.44 (0.09-1.77) | 0.27 | 0.63 (0.11-3.09) | 0.58 |
| >21 days | 0.78 (0.14-3.81) | 0.77 | 0.67 (0.10-4.00) | 0.67 |

For the association of nCoV IgG at discharge with outcome, age, sex, smoking, education, comorbidity, antiviral, and length of hospital stay were adjusted. For the association of sex, antiviral, and length of hospital stay with outcome, the aforementioned variables were all included in the model. For the association of education with outcome, the aforementioned variables except for comorbidity were included. For the association of smoking with outcome, the aforementioned variables except for comorbidity, IgG at discharge and length of hospital stay were included. For the association of comorbidity with outcome, the aforementioned variables except for IgG at discharge and length of hospital stay were included. For the association of age with outcome, only sex, cigarette smoking and education were adjusted. OR (95% CI) for age indicates the risk of outcome per 10-year age increase. OR=odds ratio. 95%CI=95% confidence interval.
